# Epidemiological and clinical features of COVID-19 patients with and without pneumonia in Beijing, China

**DOI:** 10.1101/2020.02.28.20028068

**Authors:** Penghui Yang, Yibo Ding, Zhe Xu, Rui Pu, Ping Li, Jin Yan, Jiluo Liu, Fanping Meng, Lei Huang, Lei Shi, Tianjun Jiang, Enqiang Qin, Min Zhao, Dawei Zhang, Peng Zhao, Lingxiang Yu, Zhaohai Wang, Zhixian Hong, Zhaohui Xiao, Qing Xi, Dexi Zhao, Peng Yu, Caizhong Zhu, Zhu Chen, Shaogeng Zhang, Junsheng Ji, Guangwen Cao, Fusheng Wang

## Abstract

**Background:** SARS-CoV-2-caused coronavirus disease (COVID-19) is posing a large casualty. The features of COVID-19 patients with and without pneumonia, SARS-CoV-2 transmissibility in asymptomatic carriers, and factors predicting disease progression remain unknown.

**Methods:** We collected information on clinical characteristics, exposure history, and laboratory examinations of all laboratory-confirmed COVID-19 patients admitted to PLA General Hospital. Cox regression analysis was applied to identify prognostic factors. The last follow-up was February 18, 2020.

**Results:** We characterized 55 consecutive COVID-19 patients. The mean incubation was 8.42 (95% confidence interval [CI], 6.55-10.29) days. The mean SARS-CoV-2-positive duration from first positive test to conversion was 9.71 (95%CI, 8.21-11.22) days. COVID-19 course was approximately 2 weeks. Asymptomatic carriers might transmit SARS-CoV-2. Compared to patients without pneumonia, those with pneumonia were 15 years older and had a higher rate of hypertension, higher frequencies of having a fever and cough, and higher levels of interleukin-6 (14.61 vs. 8.06pg/mL, P=0.040), B lymphocyte proportion (13.0% vs.10.0%, P=0.024), low account (<190/µL) of CD8^+^ T cells (33.3% vs. 0, P=0.019). Multivariate Cox regression analysis indicated that circulating interleukin-6 and lactate independently predicted COVID-19 progression, with a hazard ratio (95%CI) of 1.052 (1.000-1.107) and 1.082 (1.013-1.155), respectively. During disease course, T lymphocytes were generally lower, neutrophils higher, in pneumonia patients than in pneumonia-free patients. CD8^+^ lymphocytes did not increase at the 20^th^ days after illness onset.

**Conclusion:** The epidemiological features are important for COVID-19 prophylaxis. Circulating interleukin-6 and lactate are independent prognostic factors. CD8^+^ T cell exhaustion might be critical in the development of COVID-19.

## Introduction

Before 2002, 4 kinds of coronaviruses (CoVs, namely HCoV 229E, NL63, OC43, and HKU1) were known to infect humans, causing 10%-30% mild upper respiratory infection in adults, occasionally severe pneumonia in elders, infants, and immunodeficient persons.^1^ During 2002-2003, severe acute respiratory syndrome CoV (SARS-CoV) infected about 8000 persons globally, with a case fatality of 9.5%.^2^ During 2012-2015, Middle East respiratory syndrome CoV (MERS-CoV) infected about 2500 persons globally, with a case fatality rate of 35%.^1,2^ Since December 2019, an outbreak of a novel coronavirus disease (COVID-19) caused by SARS-CoV-2 had emerged in Wuhan, Hubei province, China.^3,4^ The clusters of cases were firstly reported to be related to exposure to the Huanan Seafood Market in Wuhan. Epidemiologic data indicated clear evidence of person-to-person transmission, and the transmission led to nosocomial infection.^5-7^ The case number increased quickly in Wuhan and posed a large casualty. Up to February 24, 2020, a total of 46607 confirmed cases had been identified in Wuhan, with an estimated case fatality of 4.26% (1987 deaths). SARS-CoV-2 rapidly spread to other parts of Hubei province, other parts of China, and 29 countries, resulting in 17680 cases and 508 deaths (estimated fatality: 2.87%), 12975 cases and 100 deaths (0.77%), and 2105 cases and 24 deaths (1.14%), respectively (up to February 24, 2020). The transmissibility of SARS-CoV-2 is higher and the fatality is lower compared to SARS-CoV, although the two CoVs share the receptor-human angiotensin converting enzyme II (ACE2).^8-10^ The etiological, epidemiological, and clinical characteristics of COVID-19 epidemic in Wuhan have been investigated since the disease outbreak.^3,5-7,10-14^ The occurrences of COVID-19 cases outside Wuhan were reported.^15-19^ The difference in the fatality rates indicates different epidemic patterns among the outbreak zone and outside areas. As COVID-19 is being rapidly exported to a growing number of countries, its epidemiological and clinical features outside the outbreak zone are important for the development of prophylactic options.

Currently, COVID-19 cases reported are all patients with pneumonia, rather than those without pneumonia. Furthermore, the disease course of COVID-19, the duration of viral existence, and the transmissibility of SARS-CoV-2 in asymptomatic carriers remain unknown. Here, we report a series of COVID-19 cases admitted into an infectious disease hospital that is responsible for quarantine and treatment of COVID-19 assigned by the government in Beijing, China.

## Patients and Methods

### Enrollment and diagnosis

This case series was approved by the institutional ethics board of Fifth Medical Center of PLA General Hospital, with oral informed consents. All consecutive patients with COVID-19 admitted to Fifth Medical Center of PLA General Hospital from December 27, 2019 to February 18, 2020 were enrolled. Patients with suspected COVID-19 were admitted and quarantined. Respiratory samples including throat swab samples were collected and detected for the presence of SARS-CoV-2 using a quantitative reverse transcription-PCR (qRT-PCR).^4-6^ Epidemiological information was obtained from person-to-person interview for exposure history in the past 2-3 weeks. The date of exposure is referred to the date of being Wuhan, contacting another patient from Wuhan, and visiting the related hospital(s). Incubation duration was calculated in the cases with clear cutting time points of exposure and illness onset. Clinical, laboratory, and radiographic characteristics as well as therapeutic activities and outcomes were obtained from electronic medical records. Symptoms, signs, laboratory values, chest computerized tomography scan, and treatment options during the hospital stay were all documented. COVID-19 was re-diagnosed according to the Protocol for the Diagnosis and Treatment of COVID-19 (Version 6th), National Health Commission of the People’s Republic of China. Briefly, the diagnosis of COVID-19 was confirmed if the patient’s respiratory sample was positive for SARS-CoV-2, as examined by qRT-PCR or sequencing. All diagnosed COVID-19 patients were classified as mild, common, severe, and extremely severe types. Mild type was defined if COVID-19 patient had mild symptoms but did not have pneumonia. Common type was identified if COVID-19 patient had fever, respiratory symptoms, and radiographic pneumonia. Severe type was diagnosed if COVID-19 patient with pneumonia met one of the following criteria: respiratory rate ≥30 times/min, resting oxygen saturation ≤93%, and arterial partial pressure of oxygen (PaO2)/fraction of inspired oxygen (FiO_2_) ≤300mmHg. Extremely severe type met one of the following criteria: respiratory failure that needs mechanical ventilation, shock, and combined organ failure that need to be treated in intensive care unit. Patients were discharged from hospital if they met the criteria: temperature normalized for at least 3 days, apparent improvement in respiratory symptoms, absorption of lung inflammation, and negative SARS-CoV-2 genomic RNA tested for twice at 1 day interval. The final date of follow-up was February 18, 2020.

### Statistical Analysis

Categorical variables were compared using the χ^2^ test or the Fisher exact test. Continuous variables were described using mean, median, and interquartile range (IQR) values and compared using independent group *t* tests or Mann-Whitney U test. The Cox proportional hazard model was applied to calculate the hazard ratio (HR) and 95% confidence interval (CI). Stepwise backward multivariate Cox regression analysis was performed to determine factors independently predicting the progression of COVID-19. Differences in the daily medians of laboratory parameters between patients with and without pneumonia in the 20 consecutive days were evaluated using Generalized Estimating Equations (GEE). Above statistical analyses were two-sided and performed using SPSS (Statistical Package for the Social Sciences) version 21.0 software. The dynamic diagrams and the trend of each laboratory parameter fitted by local polynomial regression (LOESS) were generated by R software (version 3.6.2). A *P* value of <0.05 was considered significant.

## Results

### Epidemiological characteristics

A total of 55 consecutive patients with laboratory-confirmed COVID-19 were enrolled in this study. Of those, 31 (56.4%) ever traveled to Wuhan or came from Wuhan within 14 days before the onset (direct exposure) and 19 (34.5%) had an indirect exposure history within 14 days before illness onset. Of the 19 cases, 14 ever hosted their sick relatives from Wuhan, 2 ate with late-diagnosed friends from Wuhan, 2 shared a railway carriage with a COVID-19 patient, and 1 attended a meeting with a late-diagnosed colleague. Confirmed case had been increasing since January 10, reached the peak on January 22, 2020, and then declined gradually. Ten days after the Wuhan quarantine on January 23, 2020, the curve touched the bottom (Figure 1). Incubation period was calculated using the data of 31 cases with clear cutting time points of exposure and illness onset. The mean incubation period of COVID-19 was 8.42 (95% CI, 6.55-10.29) days in all the cases. The mean incubation was 9.06 (95% CI, 6.11-12.00) days for patients without pneumonia and 7.54 (95% CI, 5.29-9.79) days for patients with pneumonia.

**Figure 1.**
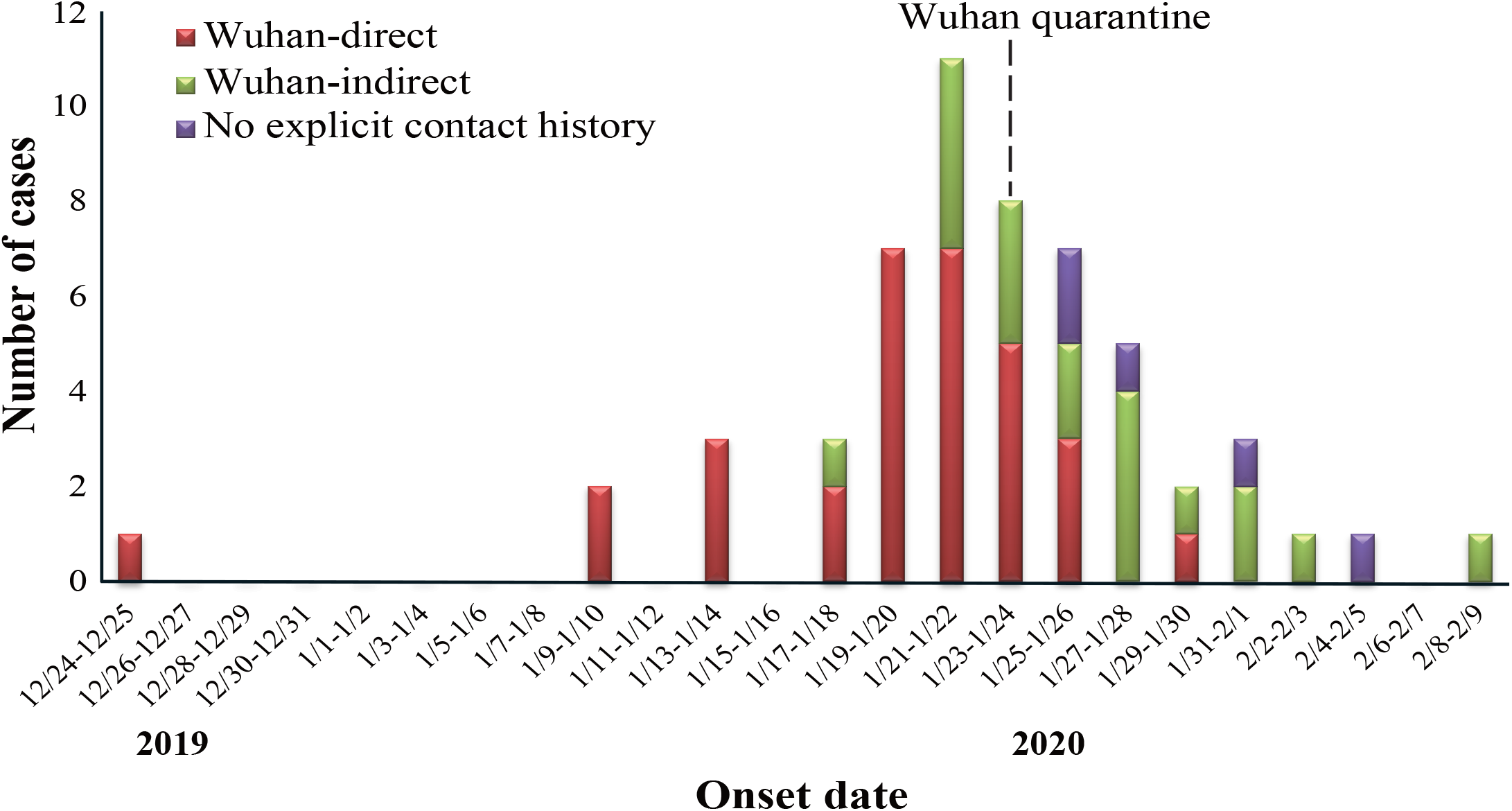
Consecutive COVID-19 patients admitted to the study hospital. Wuhan-direct, the patients who traveled to Wuhan or came from Wuhan within 14 days before illness onset; Wuhan-indirect, the patients who ever exposed to a late - diagnosed patient from Wuhan within 14 days before illness onset; and no explicit contact history, the patients did not have the above 2 situations.

Of the 55 patients, 32 had regular SARS-CoV-2 examination data. Of the 32 patients, one co-infected with human immunodeficiency virus (HIV) and hepatitis B virus (HBV) and three cases whose samples were intermittently positive were excluded. The mean (95% CI) SARS-CoV-2-positive duration from the first SARS-CoV-2-positive test to RT-PCR conversion in the remaining 28 cases was 9.71 (8.21-11.22) days. This duration was 9.24 (7.10-11.37) days for COVID-19 patients without pneumonia (n=17) and 10.45 (8.10-12.81) days for those with pneumonia (n=11).

Of the 5 remaining cases without an approved exposure history, 2 had unusual exposure. Patient A, who was long-term bed-ridden, was infected with SARS-CoV-2. She contacted only 4 healthy family members within 2 weeks before illness onset. The members did not have direct or indirect exposure history. She was shortly carried into the Emergency Department of a nearby hospital to replace her gastric tube and highly suspected to be infected there. Patient B, she and her family members did not have any exposure history within past a month. Her sister and sister’s husband from Wuhan visited her family and lived in the same building in a remote suburb of Beijing. Six days later, she became ill and was finally diagnosed as COVID-19. During the entire course, her sister’ family members were not ill or even discomfort, indicating that asymptomatic carriers transmit SARS-CoV-2.

### Clinical and laboratory parameters of COVID-19 patients with and without pneumonia at the admission

The median age was 44.0 years (IQR, 34.0-54.0; range, 3-85 years), and 22 (40.0%) were women. Major symptoms at illness onset were fever (85.5%), cough (50.9%), fatigue (27.3%), myalgia (20.0%), and sputum production (20.0%). Of the 55 patients, 26 (47.3%) had at least 1 coexisting health conditions. Hypertension (11 [20.0%]) and diabetes (6 [10.9%]) were the most common ones.

The numbers of mild, common, severe, and extremely severe patients identified at the admission were 21, 20, 13, and 1, respectively. Common, severe, and extremely severe types were combined as COVID-19 patients with pneumonia. Compared with patients without pneumonia, those with pneumonia were 15 years older and had a higher rate of hypertension, having a fever, and cough (Table 1). Compared with COVID-19 patients without pneumonia, those with pneumonia had higher levels of interleukin-6, B lymphocyte proportion, erythrocyte sedimentation rate, and very low count (<190/µL) of CD8^+^ T cells (Table 2).

**Table 1.** Baseline information of COVID-19 patients enrolled in this study

|  | All enrolled patients<br>(n=55) | Patients without<br>pneumonia (n=21) | Patients with<br>pneumonia (n=34) | P |
| --- | --- | --- | --- | --- |
| Age, Median (IQR) – yrs | 44(34.00-54.00) | 35(22.50-44.50) | 50(39.00-63.25) | <0.001 |
| Gender |  |  |  | 0.821 |
| Male | 33(60.0) | 13(61.9) | 20(58.8) |  |
| Female | 22(40.0) | 8(38.1) | 14(41.2) |  |
| BMI, Median (IQR) | 24.85(22.86-26.79) | 26.12(22.86-27.68) | 24.54(22.89-26.27) | 0.398 |
| Exposure |  |  |  | 0.201 |
| No explicit contact | 5(9.1) | 0 | 5(14.7) |  |
| Wuhan-direct | 31(56.4) | 14(66.7) | 17(50.0) |  |
| Wuhan-indirect | 19(34.5) | 7(33.3) | 12(35.3) |  |
| Contact history |  |  |  | 0.226 |
| No | 31(56.4) | 14(66.7) | 17(50.0) |  |
| Yes | 24(43.6) | 7(33.3) | 17(50.0) |  |
| Underlying diseases |  |  |  |  |
| Hypertension | 11(20.0) | 0 | 11(32.4) | 0.004 |
| Diabetes | 6(10.9) | 2(9.5) | 4(11.8) | 1.000 |
| AIDS | 1(1.8) | 0 | 1(2.9) | 1.000 |
| Chronic liver disease | 1(1.8) | 0 | 1(2.9) | 1.000 |
| Appendectomy | 2(3.6) | 0 | 2(5.9) | 0.519 |
| Lung disease | 3(5.5) | 1(4.8) | 2(5.9) | 1.000 |
| Fracture | 2(3.6) | 1(4.8) | 1(2.9) | 1.000 |
| Tonsillectomy | 1(1.8) | 0 | 1(2.9) | 1.000 |
| Cerebral infarction | 1(1.8) | 0 | 1(2.9) | 1.000 |
| Asthma | 1(1.8) | 0 | 1(2.9) | 1.000 |
| Stone | 3(5.5) | 0 | 3(8.8) | 0.279 |
| Gallbladder surgery | 2(3.6) | 0 | 2(5.9) | 0.519 |
| Intestinal polypectomy | 1(1.8) | 0 | 1(2.9) | 1.000 |
| Cesarean section | 2(3.6) | 0 | 2(5.9) | 0.519 |
| Cervical spondylosis | 1(1.8) | 0 | 1(2.9) | 1.000 |
| Thyroid disease | 2(3.6) | 1(4.8) | 1(2.9) | 1.000 |
| Main symptoms |  |  |  |  |
| Fever | 47(85.5) | 15(71.4) | 32(94.1) | 0.043 |
| Cough | 28(50.9) | 7(33.3) | 21(61.8) | 0.040 |
| Headache | 6(10.9) | 2(9.5) | 4(11.8) | 1.000 |
| Sore throat | 9(16.4) | 5(23.8) | 4(11.8) | 0.279 |
| Myalgia | 11(20.0) | 4(19.0) | 7(20.6) | 1.000 |
| Dizziness | 9(16.4) | 6(28.6) | 3(8.8) | 0.071 |
| Sputum production | 11(20.0) | 4(19.0) | 7(20.6) | 1.000 |
| Diarrhea | 2(5.5) | 0 | 2(5.9) | 0.519 |
| Chest tightness | 3(5.5) | 0 | 3(8.8) | 0.279 |
| Dyspnoea | 5(9.1) | 0 | 5(14.70) | 0.144 |
| Fatigue | 15(27.3) | 2(9.5) | 13(38.2) | 0.020 |
| Stuffy nose | 1(1.8) | 1(4.8) | 0 | 0.382 |
| Rash | 2(3.6) | 1(4.8) | 1(2.9) | 1.000 |
| Highest temperature, °C |  |  |  | 0.855 |
| 37.3–38.0 | 19/48(39.6) | 6/17(35.3) | 13/31(41.9) |  |
| 38.1–39.0 | 20/48(41.7) | 7/17(41.2) | 13/31(41.9) |  |
| >39.0 | 9/48(18.8) | 4/17(23.5) | 5/31(16.1) |  |
| Fever duration, Median | 5.5(2.00-8.00) | 5(2.75-6.25) | 6(2.00-9.75) | 0.072 |
| (IQR), day |  |  |  |  |
| Systolic pressure, Median | 124(117.75-133.50) | 125(120.00-133.50) | 121(116.50-134.00) | 0.823 |
| (IQR), mmHg |  |  |  |  |

Data are median (IQR), n (%), or n/N (%), where N is the total number of patients with available data. P values compare patients with and without pneumonia using $\chi^2$ test, Fisher's exact test, or student *t* test.

**Table 2.** Comparison of laboratory findings at the admission between COVID-19 patients with and without pneumonia

|  | All enrolled patients<br>(n=55) | Patients without<br>pneumonia (n=21) | Patients with pneumonia<br>(n=34) | P |
| --- | --- | --- | --- | --- |
| Blood Glucose, mmol/L | 5.30(4.90-6.70) | 5.20(4.80-5.70) | 5.50(5.00-7.15) | 0.554 |
| <3.9 | 1/48(2.1) | 1/19(5.3) | 0 | 0.106 |
| 3.9-6.1 | 33/48(68.8) | 15/19(78.9) | 18/29(62.1) |  |
| >6.1 | 14/48(29.2) | 3/19(15.8) | 11/29(37.9) |  |
| Blood Lipid, mmol/L | 1.17(0.82-1.51) | 1.48(0.82-1.60) | 1.13(0.82-1.32) | 0.893 |
| PH | 7.46(7.44-7.47) | 7.46(7.44-7.47) | 7.46(7.43-7.48) | 0.773 |
| 7.35-7.45 | 13/34(38.2) | 3/7(42.9) | 10/27(37.0) | 1.000 |
| >7.45 | 21/34(61.8) | 4/7(57.1) | 17/27(63.0) |  |
| PaO<sub>2</sub>, mmHg | 78.00(86.50-127.75) | 92.00(80.00-148.00) | 86.00(75.00-115.00) | 0.428 |
| <80 | 10/34(29.4) | 1/7(14.3) | 9/27(33.3) | 0.660 |
| 80-100 | 12/34(35.3) | 3/7(42.9) | 9/27(33.3) |  |
| >100 | 12/34(35.3) | 3/7(42.9) | 9/27(33.3) |  |
| PaCO<sub>2</sub>, mmHg | 37.00(34.75-40.00) | 39.00(35.00-41.00) | 37.00(34.00-38.00) | 0.436 |
| <35 | 8/34(23.5) | 1/7(14.3) | 7/27(25.9) | 0.723 |
| 35-45 | 25/34(73.5) | 6/7(85.7) | 19/27(70.4) |  |
| >45 | 1/34(2.9) | 0 | 1/27(3.7) |  |
| SaO<sub>2</sub>, mmHg | 97.00(96.00-99.00) | 96.50(96.00-99.00) | 97.00(95.00-98.50) | 0.539 |
| ABE, mmol/L | 1.75(0.40-2.40) | 1.65(-0.53-3.08) | 1.75(0.40-2.23) | 0.995 |
| <-3 | 1/22(4.5) | 0 | 1/18(5.6) | 1.000 |
| -3-3 | 17/22(77.3) | 3/4(75.0) | 14/18(77.8) |  |
| >3 | 4/22(18.2) | 1/4(25.0) | 3/18(16.7) |  |
| Leukocyte, 10<sup>9</sup>/L | 4.64(3.69-5.95) | 5.17(4.11-6.15) | 4.31(3.41-6.12) | 0.340 |
| <3.97 | 18(32.7) | 4(19.0) | 14(41.2) | 0.139 |
| 3.97-9.15 | 36(65.5) | 17(81.0) | 19(55.9) |  |
| >9.15 | 1(1.8) | 0 | 1(2.9) |  |
| Neutrophils, % | 59.75(52.45-67.70) | 57.30(52.80-67.10) | 62.40(50.75-67.90) | 0.267 |
| <50 | 10/54(18.5) | 3(14.3) | 7/33(21.2) | 0.567 |
| 50-70 | 34/54(63.0) | 15(71.4) | 19/33(57.6) |  |
| >70 | 10/54(18.5) | 3(14.3) | 7/33(21.2) |  |
| Lymphocyte, % | 30.40(21.78-38.53) | 32.80(23.35-36.30) | 28.90(20.60-38.85) | 0.387 |
| <20 | 11/54(20.4) | 3(14.3) | 8/33(24.2) | 0.640 |
| 20-40 | 32/54(59.3) | 14(66.7) | 18/33(54.5) |  |
| >40 | 11/54(20.4) | 4(19.0) | 7/33(21.2) |  |
| C reactive protein, mg/L | 7.95(1.79-25.06) | 7.10(1.20-29.50) | 9.30(3.60-21.60) | 0.378 |
| 0.06-88.2 | 51/52(98.1) | 19/19(100.0) | 32/33(97.0) | 1.000 |
| >88.2 | 1/52(1.9) | 0 | 1/33(3.0) |  |
| IL-6, pg/mL | 11.26(5.39-24.98) | 8.06(5.38-15.61) | 14.61(5.39-27.31) | 0.040 |
| 0-7 | 21/52(40.4) | 9/19(47.4) | 12/33(36.4) | 0.436 |
| >7 | 31/52(40.4) | 10/19(52.6) | 21/33(63.6) |  |
| Procalcitonin, ng/mL | 0.04(0.03-0.07) | 0.04(0.03-0.06) | 0.05(0.03-0.08) | 0.168 |
| 0-0.5 | 52/53(98.1) | 20/20(100.0) | 32/33(97.0) | 1.000 |
| >0.5 | 1/53(1.9) | 0 | 1/33(3.0) |  |
| ESR, mm/60min | 10.00(2.10-31.00) | 6.00(2.03-13.00) | 17.00(5.00-39.50) | 0.006 |
| 0-15 | 33/53(62.3) | 17/20(85.0) | 16/33(48.5) | 0.008 |
| >15 | 20/53(37.7) | 3/20(15.0) | 17/33(51.5) |  |
| Lactate, mmol/L | 2.15(1.59-4.10) | 2.08(1.68-5.06) | 2.24(1.57-3.79) | 0.736 |
| 0.6-2.2 | 23/45(51.1) | 8/14(57.1) | 15/31(48.4) | 0.586 |
| >2.2 | 22/45(48.9) | 6/14(42.9) | 16/31(51.6) |  |
| Lymphocytes, /μL | 1100.00(771.00-1374.00) | 1113.00(883.00-1406.00) | 1097.50(769.50-1371.50) | 0.663 |
| <800 | 12/41(29.3) | 3/13(23.1) | 9/28(32.1) | 0.719 |
| 800-4000 | 29/41(70.7) | 10/13(76.9) | 19/28(67.9) |  |
| T lymphocyte, % | 69.00(62.50-77.00) | 71.00(62.50-77.00) | 68.00(62.00-77.00) | 0.720 |
| <55 | 6/37(16.2) | 1/10(10.0) | 5/27(18.5) | 0.553 |
| 55-84 | 28/37(75.7) | 9/10(90.0) | 19/27(70.4) |  |
| >84 | 3/37(8.1) | 0 | 3/27(11.1) |  |
| T lymphocyte count, /μL | 785.00(488.00-994.75) | 802.00(519.00-931.00) | 768.00(408.00-1024.00) | 0.919 |
| <690 | 14/38(36.8) | 4/11(36.4) | 10/27(37.0) | 1.000 |
| 690-2540 | 24/38(63.2) | 7/11(63.6) | 17/27(63.0) |  |
| CD4<sup>+</sup> lymphocyte, % | 38.00(29.00-42.00) | 39.50(26.75-41.25) | 36.00(29.00-44.00) | 0.766 |
| <31 | 10/37(27.0) | 3/10(30.0) | 7/27(25.9) | 1.000 |
| 31-60 | 27/37(73.0) | 7/10(70.0) | 20/27(74.1) |  |
| CD4<sup>+</sup> lymphocyte count, /μL | 425.50(265.00-543.00) | 425.00(285.00-619.00) | 435.00(262.00-540.00) | 0.185 |
| <410 | 18/40(45.0) | 5/13(38.5) | 13/27(48.1) | 0.564 |
| 410-1590 | 22/40(55.0) | 8/13(61.5) | 14/27(51.9) |  |
| CD8<sup>+</sup> lymphocyte, % | 28.00(23.00-35.25) | 28.00(24.00-35.00) | 30.00(23.00-36.00) | 0.440 |
| <13 | 3/38(7.9) | 0 | 3/27(11.1) | 0.416 |
| 13-41 | 33/38(86.8) | 10/11(90.9) | 23/27(85.2) |  |
| >41 | 2/38(5.3) | 1/11(9.1) | 1/27(3.7) |  |
| CD8<sup>+</sup> lymphocyte count, /μL | 337.00(200.25-453.75) | 366.00(269.00-421.50) | 318.00(144.00-477.00) | 0.307 |
| <190 | 9/40(22.5) | 0 | 9/27(33.3) | 0.019 |
| 190-1140 | 31/40(77.5) | 13/13(100.0) | 18/27(66.7) |  |
| B lymphocyte, % | 12.00(9.50-16.50) | 10.00(9.50-12.25) | 13.00(9.00-21.00) | 0.024 |
| 6-25 | 32/36(88.9) | 10/10(100.0) | 22/26(84.6) | 0.559 |
| >25 | 4/36(11.1) | 0 | 4/26(15.4) |  |
| B lymphocyte count, /μL | 126.00(90.50-185.50) | 120.50(98.00-132.00) | 126.00(89.00-190.00) | 0.403 |
| <90 | 9/37(24.3) | 2/10(20.0) | 7/27(25.9) | 1.000 |
| 90-660 | 28/37(75.7) | 8/10(80.0) | 20/27(74.1) |  |
| NK lymphocyte, % | 16.00(9.50-24.00) | 18.00(11.75-35.25) | 16.00(8.00-23.00) | 0.252 |
| 5-27 | 30/36(83.3) | 7/10(70.0) | 23/26(88.5) | 0.317 |
| >27 | 6/36(16.7) | 3/10(30.0) | 3/26(11.5) |  |
| NK lymphocyte count, /μL | 162.00(74.50-226.50) | 159.00(134.25-264.50) | 167.00(68.00-221.00) | 0.754 |
| <90 | 10/37(27.0) | 1/10(10.0) | 9/27(33.3) | 0.229 |
| 90-590 | 27/37(73.0) | 9/10(90.0) | 18/27(66.7) |  |
| CD4/CD8 | 1.19(0.91-1.60) | 1.34(1.00-1.68) | 1.15(0.85-1.61) | 0.350 |
| <0.68 | 4/41(9.8) | 2/14(14.3) | 2/27(7.4) | 0.725 |
| 0.68-2.47 | 32/41(78.0) | 11/14(78.6) | 21/27(77.8) |  |
| >2.47 | 5/41(12.2) | 1/14(7.1) | 4/27(14.8) |  |
| NLR | 1.97(1.39-2.97) | 1.75(1.48-2.75) | 2.16(1.31-3.26) | 0.259 |

Abbreviations: PH, hydrogen ion concentration; PaO<sub>2</sub>, partial pressure of oxygen; PaCO<sub>2</sub>, partial pressure of carbon dioxide in artery; SaO<sub>2</sub>, arterial oxygen saturation; IL-6, interleukin-6; ABE, actual base excess; ESR, Erythrocyte sedimentation rate; NLR, Neutrophils/Lymphocytes.
Data are median (IQR), n (%), or n/N (%), where N is the total number of patients with available data. P values compare patients with and without pneumonia using $\chi^2$ test, Fisher's exact test, or student *t* test.

### Disease course

Of the 55 patients, 52 received antiviral treatment, mostly with lopinavir/ritonavir and or arbidol. The mean duration from onset to admission was 5.81(95% CI, 4.95-6.68) days in 54 patients (except the HIV case). The mean duration was 4.86 (3.46-6.25) in pneumonia-free patients and 6.42 (5.23-7.53) in pneumonia patients. The mean duration from illness onset to discharge was 18.57 (16.07-21.08) days in the 28 patients. The mean duration was 17.76 (14.45-21.08) days in pneumonia-free patients and 19.82 (15.39-24.24) days in pneumonia patients. According to the discharge criteria, disease course of COVID-19 was approximately 2 weeks, which is in accordance with another group of patients without antiviral treatment.^18^

### Factors predicting progressive disease

During hospital stay, 1 case with common-type COVID-19 progressed to be severe COVID-19, 4 severe patients progressed to be extremely severe COVID-19. Of the 4 patients, 2 died of COVID-19. Another 2 cases have been staying in the hospital near a month due to the development of extra-respiratory signs. The 7 cases were categorized as patients with progressive disease. Univariate Cox analysis indicated that hypertension, circulating IL-6, and circulating lactate significantly predicted the progression. Multivariate Cox regression analysis indicated that circulating IL-6 and lactate independently predicted COVID-19 progression (Table 3).

**Table 3.** Cox regression analyses for factors predicting the progression of COVID-19

| Covariate | Univariate analysis | P | Multivariate analysis |  |
| --- | --- | --- | --- | --- |
|  | HR (95% CI) |  | HR (95% CI) | P |
| Age, yrs | 1.040(0.991-1.092) | 0.110 |  |  |
| Hypertension | 10.182(1.125-92.186) | **0.039** |  |  |
| Fever | 1.069(0.117-9.799) | 0.953 |  |  |
| Cough | 1.989(0.360-10.993) | 0.430 |  |  |
| C reactive protein, mg/L | 1.025(0.992-1.059) | 0.140 |  |  |
| T lymphocyte count, /μL | 1.000(0.996-1.004) | 0.868 | | |
| CD4<sup>+</sup> lymphocyte count, /μL | 1.000(0.996-1.005) | 0.927 | | |
| CD8<sup>+</sup> lymphocyte count, /μL | 0.999(0.993-1.005) | 0.790 | | |
| B lymphocyte count, /μL | 0.997(0.981-1.012) | 0.674 | | |
| NK lymphocyte count, /μL | 1.002(0.994-1.010) | 0.682 | | |
| IL-6, pg/ml | 1.040(1.002-1.080) | **0.037** | 1.052(1.000-1.107) | **0.048** |
| Lactate, mmol/L | 1.072(1.010-1.138) | **0.022** | 1.082(1.013-1.155) | **0.018** |
| NLR | 1.066(0.891-1.276) | 0.482 |  |  |
| CD4<sup>+</sup>/total T lymphocyte | 1.798(0.022-148.008) | 0.794 |  |  |
| CD8<sup>+</sup>/total T lymphocyte | 0.787(0.008-73.150) | 0.918 |  |  |

Abbreviations: IL 6, interleukin-6; NLR, Neutrophils/Lymphocytes; HR, hazard ratio; CI, confidence interval.

We then evaluated the dynamics of clinical and laboratory parameters during hospital stay between COVID-19 patients with pneumonia and those without pneumonia and re-categorized the patient with progressive disease. Of the parameters evaluated, 6 showed different trends between COVID-19 patients with and without pneumonia (P<0.001 for each, GEE). The daily medians of neutrophil proportions and neutrophil/lymphocyte ratio (NLR) were higher in pneumonia patients than in pneumonia-free patients, which were quite in contrast to lymphocyte proportion, T lymphocyte count, CD4^+^ T lymphocyte count, and CD8^+^ T lymphocyte count. At the 20^th^ day after illness onset, the dynamic curves of neutrophil proportion, NLR, lymphocyte proportion, and CD4^+^ T lymphocyte count tended to meet (P>0.05), except those of CD8^+^ T lymphocytes and T lymphocyte counts (P<0.05) (Figure 2).

**Figure 2.**
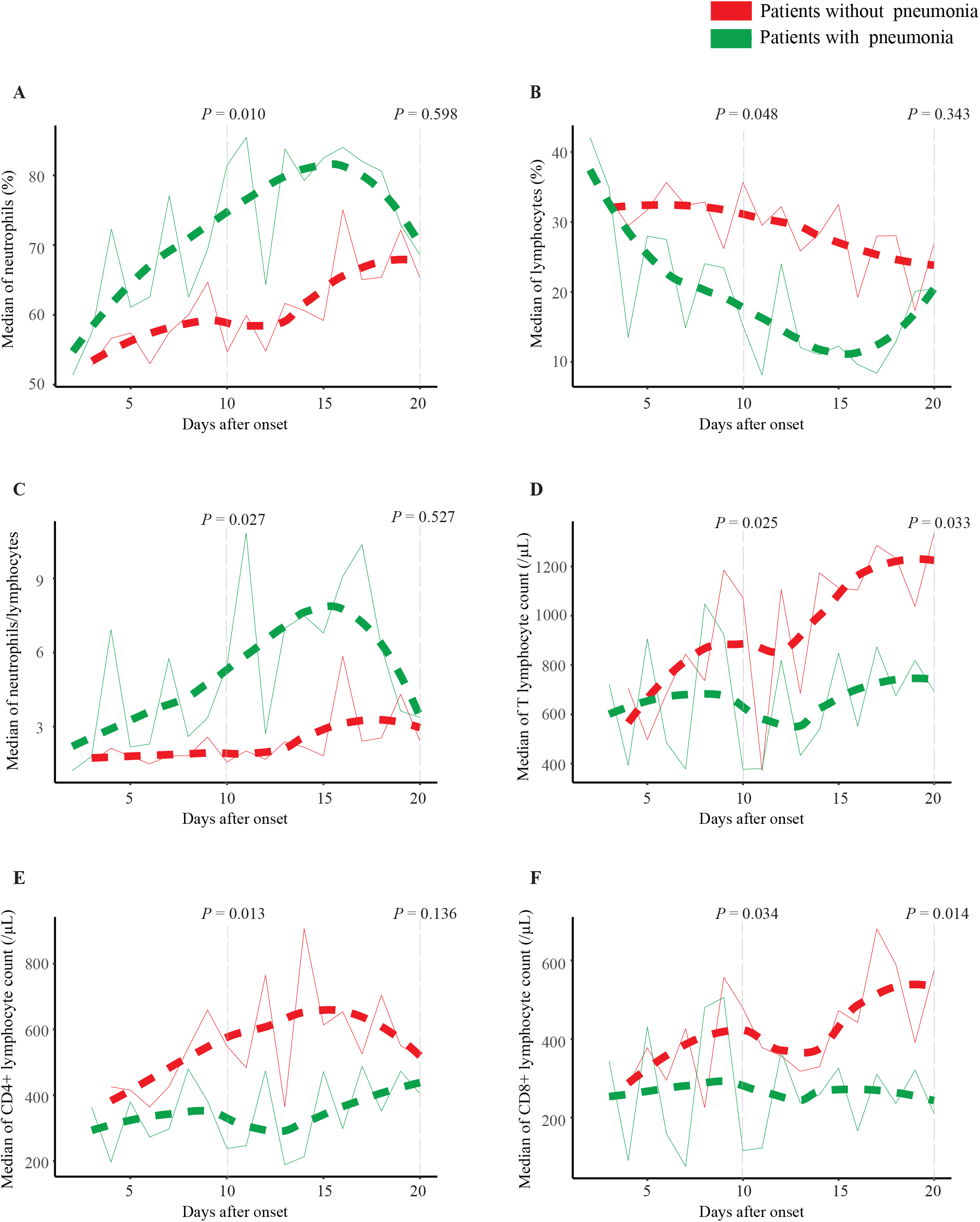
The dynamic curves of major laboratory findings between COVID-19 patients with pneumonia and those without pneumonia. Daily medians of laboratory parameters within the first 20 days were applied to construct the dynamic curves. P value indicates the difference between laboratory parameters at the 10^th^ and 20^th^ day since illness onset (Mann-Whitney U test).

## Discussion

The epidemiological and clinical features of COVID-19 in Beijing might represent the features of COVID-19 outside the outbreak zone. Most patients had clear exposure history, indicating that SARS-CoV-2 transmits predominantly via respiratory droplet. As SARS-CoV-2 shares its receptor with SARS-CoV,^6^ transmission dynamics of SARS-CoV could be referenced. Person-to-person transmission of SARS-CoV occurs mainly in the second week of illness when the patient’s upper airway viral load peaks on day 10 of illness.^20^ We found that SARS-CoV-2 was mostly transmitted via contacting symptomatic patients; however, asymptomatic carriers might transmit SARS-CoV-2 to healthy recipients. This might be one of the reasons why SARS-CoV-2 is more transmissible than is SARS-CoV. Thus, personnel quarantine, avoiding any kind of party or gathering, and avoiding sharing public facility are necessary during COVID-19 epidemic. Elders with underlying medical conditions including hypertension often resulted in serious consequences. This observation is quite in accordance with SARS and MERS.^20^ Specific prophylaxis should be given to this vulnerable population.

We revealed that the mean incubation of SARS-CoV-2 infection was 8.42 days. A previous study carried out in Wuhan indicated that the mean incubation period was 5.2 days.^5^ Early patients identified in the outbreak zone were mostly with severe pneumonia and might be exposed to higher viral concentration in closed spaces like the affected hospitals. Furthermore, it is hard to identify the exact time of exposure in the outbreak zone. Thus, the incubation from this study should be important in quarantining close contactors outside the outbreak zone.

The durations of COVID-19 course and SARS-CoV-2-positive course are not reported previously. We determined that the mean duration from onset to discharge was 18.57 days. According to the discharge criteria, COVID-19 course is approximately 2 weeks, which is quite in consistent with SARS.^21^ The mean SARS-CoV-2-positive duration from the first positive test to RT-PCR conversion was 9.71 days. SARS-CoV-2 might be present in patients for around 18 days by adding the mean incubation of 8.42 days. This duration seems to be shorter than SARS.^22^ A just finished clinical study in Shanghai did not show any therapeutic effects of lopinavir/ritonavir and abidol on accelerating the clearance SARS-CoV-2.^23^ Thus, the current antiviral regimen could not alter the course of COVID-19.

To our knowledge, this was the first study to characterize the differences in the clinical features and laboratory findings between COVID-19 patients with and without pneumonia. Unlike those without pneumonia, one-third of COVID-19 patients with pneumonia had very low account (<190/µL) of CD8^+^ T cells. In addition, serum level of IL-6 was higher in COVID-19 patients with pneumonia than in those without pneumonia (Table 2). These data suggest that CD8^+^ T cell exhaustion and IL-6-based inflammation might play an important role in the development of COVID-19.

It is critical to identify COVID-19 patients who are most likely to have poor outcomes so that prevention and treatment efforts can be focused.^24^ Here, we reported that serum IL-6 and lactate at the admission independently predicted poor outcomes of COVID-19 patients, after the adjustment with other significant factors in the univariate Cox analysis (Table 3). IL-6 is capable of inhibiting the T cell-mediated immunity, and high level of IL-6 in the acute stage is associated with lung lesions in SARS patients.^25^ Lactate is a natural by-product of aerobic glycolysis. Efferocytosis can promote glucose uptake and lactate release.^26^ Lactate promotes the switch of CD4^+^ T cells to an IL-17^+^ subset and impairs the cytolytic capacity of CD8^+^ T cells.^27^ Thus, IL-6 might bridge SARS-CoV-2 infection and alveolar cell injury via inducing tissue-damaging inflammation. The inflammation increases efferocytosis-mediated aerobic glycolysis, facilitating the release of lactate. Increased lactate in turn impairs the CD8^+^ T cell-mediated antiviral immunity.

For the first time, the dynamic changes of laboratory parameters of COVID-19 patients with and without pneumonia were evaluated during hospital stay. Neutrophil proportion and NLRs were higher in pneumonia patients than in pneumonia-free patients, indicating systemic inflammation is apparent. Lymphocyte proportion and CD4^+^ T lymphocyte counts were lower in pneumonia patients than in pneumonia-free, and the two curves met at the 20th day, indicating B cells might recover rapidly. Surprisingly, the lower levels of CD8^+^ cells in pneumonia patients did not increase at the 20th day (Figure 2), indicating that recovery of CD8^+^ T cell-mediated immunity needs a longer time. Thus, the discharged patients might be susceptible to infections and other diseases. The recovery of T cell count should be monitored.

Our study has several limitations. First, the effect of antiviral treatment was not evaluated due to lack of controls. Second, the family members of patient B’s sister were not examined for SARS-CoV-2, because they left when we performed this study. Third, half patients are still hospitalized. The disease course of severe patients might be prolonged by therapeutic regimens including corticosteroids.^28^

Finally, we characterized the epidemiological features including the incubation period, time to RT-PCR conversion of SARS-CoV-2, COVID-19 course, and the transmissibility SARS-CoV-2 in asymptomatic carriers. Old age, hypertension, interleukin-6, and CD8^+^ T cell exhaustion were significantly associated with the development of pneumonia. Circulating interleukin-6 and lactate independently predicted COVID-19 progression. CD8^+^ T cell exhaustion might be important in COVID-19 progression. This study not only elucidates some mechanisms of COVID-19 progression, but also provides epidemiological evidence to prevent COVID-19 epidemic outside the outbreak zone.

## Data Availability

Original data are open.

